# Genome-wide association study in chondrocalcinosis reveals ENPP1 as a candidate therapeutic target in calcium pyrophosphate deposition disease

**DOI:** 10.1101/2024.10.10.24315203

**Authors:** Riku Takei, Ann Rosenthal, Tristan Pascart, Richard J. Reynolds, Sara K. Tedeschi, Tony R. Merriman

## Abstract

**Objective:** The genetic basis of calcium pyrophosphate deposition (CPPD) disease is largely unknown. This limits the development of therapeutic strategies. We aimed to analyze a genome-wide association study (GWAS) on a large administrative database to identify new candidate causal genes for CPPD disease.

**Methods:** We used publicly available GWAS summary statistics for chondrocalcinosis and for crystal arthropathy from the Veterans Affairs Million Veteran Program in people of African (AFR) and European (EUR) ancestry. Included were 3,004 (536 AFR and 2,468 EUR) cases for chondrocalcinosis and 3,766 (700 AFR and 3,066 EUR) cases for crystal arthropathy. Our primary analysis was in chondrocalcinosis with secondary analysis in crystal arthropathy. We tested for colocalization of chondrocalcinosis genetic association signals with genetic control of gene expression.

**Results:** There were two genome-wide significant loci for chondrocalcinosis in both AFR and EUR, both on chromosome 6 (signals within the *ENPP1* and *RNF144B* genes). Findings were supported by analysis of the crystal arthropathy cohort. Colocalization analysis of chondrocalcinosis genetic association signals with genetic control of gene expression and alternative splicing further supported *ENPP1* and *RNF144B* as candidate casual genes. At *ENPP1* the allele that increases the risk for chondrocalcinosis associated with increased *ENPP1* expression.

**Conclusion:** *ENPP1* encodes ectonucleotide pyrophosphatase / phosphodiesterase family member 1 that produces AMP and pyrophosphate, potentially contributing to the formation of calcium pyrophosphate crystals. Selective ENPP1 inhibitors developed for infectious disease and cancer could be repurposed for the treatment of chondrocalcinosis and CPPD disease.

## INTRODUCTION

The deposition of calcium pyrophosphate crystals in articular tissues characterizes calcium pyrophosphate deposition (CPPD) disease. CPPD disease is a heterogeneous crystalline arthritis that can cause acute or chronic joint symptoms and is claimed to be the most common inflammatory arthritis in people over 60 years of age^1-4^. Acute calcium pyrophosphate (CPP) crystal arthritis, historically called “pseudogout,” is the most widely-recognized form of CPPD disease. It results from CPP crystals activating the NLRP3 inflammasome, leading to IL-1β secretion and causing acute inflammatory arthritis^5^. Chondrocalcinosis, a radiographic finding that is most often due to CPPD^6-8^ (much less commonly basic calcium phosphate), is a common finding in older adults and increases in prevalence with each decade beyond age 60. In Europe and North America, the prevalence of imaging evidence of CPPD is estimated to be about 10% in middle-aged adults, depending on articular location^9^, with prevalence increasing to approximately 30% in adults over 80 years of age^10,11^. CPPD also associates with cartilage degradation and osteoarthritis, although it remains unclear whether CPPD is a cause or consequence of these conditions^12^.

There are no effective treatments for the prevention and/or dissolution of CPP crystals that would be akin to the use of urate-lowering therapy in gout, a common inflammatory arthritis also cause by deposition of crystals (monosodium urate) in joints. The pathogenesis of CPPD disease remains incompletely understood, although increased inorganic pyrophosphate (PPi) is central to the formation of CPP crystals in and around cartilage. Treatments for CPPD disease focus on the alleviation of inflammation^3^ most often with non-steroidal anti-inflammatory drugs, colchicine or prednisone; biologics inhibiting IL-1β and IL-6 have shown promise for patients with recurrent acute or chronic inflammatory CPP crystal arthritis^1,3^. The US-based Gout, Hyperuricemia and Crystal-Associated Disease Network (G-CAN) has recognized that there is considerable unmet need in CPPD disease^13^.

Identifying genetic associations with disease by genome-wide association studies (GWAS) has revolutionized understanding of disease etiology. However, despite the prevalence and impact of CPPD on the middle-aged and elderly population, no GWAS has been done to date. There is virtually no knowledge on the genetic basis of CPPD, with genetic studies limited to various candidate gene studies such as *ANKH^14,15^*, mostly because CPPD disease is not directly recorded in large biobanks*^1^*.Here we report the findings of the first GWAS for chondrocalcinosis (cartilage calcification on imaging) and for crystal arthropathy.

## METHODS

### Million Veteran Program cohorts

The overall US Department of Veterans Affairs Million Veteran Program (MVP) cohort is 91.2% male with mean age of 61.9 years^16^. There were 121,177 individuals with genetic similarity to the 1000 Genomes Project African cohort and 449,042 individuals with genetic similarity to the European cohort^16^. Given that this study includes only data from the MVP we define the ancestral groups used here as African American (AFR) and European American (EUR). We used publicly-available summary statistics for each of chondrocalcinosis (Phecode 274.21) and crystal arthropathy (Phecode 274.2) from a systematic analysis of 2068 traits in 635,969 people from the MVP^16^. The Phecodes had been extracted from Veterans Affairs electronic records^16^. Phecodes are aggregates of International Classification of Diseases, 9^th^ Revision, Clinical Modification (ICD-9-CM) and ICD-10-CM codes that have been manually reviewed and clustered to align with distinct diseases or traits^17^. Phecodes enable high-throughput phenotyping using electronic health record data and outperformed ICD-9-CM codes in representing clinical phenotypes associated with known genetic variants^18^. Numbers of cases and controls for each phenotype are in the legend of **Table 1**. Ethical approval was not required for this study as freely downloadable publicly available summary statistics were used.

**Table 1.**

|  | SNP | Position<br>Chr:Base pairs | Risk allele:<br>frequency | OR [95% CI] | *P* | Closest<br>gene |
| --- | --- | --- | --- | --- | --- | --- |
| AFR<sup>1</sup> |  |  |  |  |  |  |
| CC | rs9396861 | 6:18403902 | C:0.36 | 1.49 [1.29, 1.71] | 3.4x10<sup>-8</sup> | RNF144B |
| CC | rs11963689 | 6:131889538 | C:0.80 | 1.78 [1.47, 2.15] | 3.8x10<sup>-9</sup> | ENPP1 |
| CA | rs11963689 | 6:131889538 | C:0.80 | 1.64 [1.40, 1.93] | 2.1x10<sup>-9</sup> | ENPP1 |
| EUR<sup>1</sup> |  |  |  |  |  |  |
| CC | rs1886248 | 6:18403902 | C:0.59 | 1.43 [1.35, 1.51] | 4.0x10<sup>-33</sup> | RNF144B |
| CC | rs6939185 | 6:131818047 | G:0.60 | 1.32 [1.24, 1.40] | 3.5x10<sup>-19</sup> | ENPP1 |
| CA | rs1886248 | 6:18399163 | C:0.59 | 1.36 [1.28, 1.43] | 2.7x10<sup>-31</sup> | RNF144B |
| CA | rs766592 | 6:131810461 | G:0.60 | 1.27 [1.20, 1.34] | 1.8x10<sup>-17</sup> | ENPP1 |
| TAMA |  |  |  |  |  |  |
| CC | rs1886248 | 6:18403902 | C:0.39 | 1.43 [1.35, 1.50] | 3.6x10<sup>-40</sup> | RNF144B |
| CC | rs1409181 | 6:131828160 | G:0.47 | 1.26 [1.19, 1.33] | 2.3x10<sup>-17</sup> | ENPP1 |
| CA | rs9396861 | 6:18403902 | C:0.46 | 1.37 [1.30, 1.44] | 2.5x10<sup>-36</sup> | RNF144B |
| CA | rs1409181 | 6:131828160 | G:0.47 | 1.21 [1.27, 1.16] | 3.8x10<sup>-15</sup> | ENPP1 |

1 There were 536 AFR CC cases and 120,708 controls; 700 AFR CA cases and 120,306 controls; 2,468 EUR CC cases and 445,620 controls; 3,066 EUR CA cases and 444,490 controls. AFR, African; CC, chondrocalcinosis; CA, crystal arthropathies; EUR, European.

### Analysis of GWAS summary statistics

Genotyping had been done using a customized ThermoFisher Axiom MVP 1.0 platform and imputation done using a hybrid reference panel of the African Genome Resources Panel available at the Sanger Imputation Service, and 1000 Genomes^16^ with subsequent variant level quality control described in ref^16^. The GWAS had been performed using the Scalable and Accurate Implementation of Generalized (SAIGE) mixture model^19^ with adjustment for age, sex and the first 10 principal components of genetic variation^16^. There were no summary statistics for either phenotype provided in ref^16^ for any other population groups. We analyzed 26,879,965 SNPs with minor allele frequency (MAF) > 0.1% (in combined cases and controls) in the AFR and 13,836,906 in the EUR cohort for chondrocalcinosis and 26,879,949 SNPs in AFR and 13,836,871 in EUR for crystal arthropathy. The metadata provided in ref^16^ had removed variants with a minor allele count < 30 and imputation quality < 0.3. AFR and EUR summary statistics for each of chondrocalcinosis and crystal arthropathy were also meta-analyzed by an inverse-variance weighted random effects method using GWAMA software^20^. Locus Zoom plots were drawn using code available from ref^21^. A genetic association signal was declared when > 1 SNP at a locus was genome-wide significant (*P* < 5×10^-8^). Colocalization analysis testing for overlap between chondrocalcinosis genetic association signals and genetic control of gene expression (expression quantitative trait locus (eQTL)) and alternative splicing (splicing quantitative trait locus (sQTL)) was done using the ‘coloc’ R package^22,23^ (signals were deemed colocalized if the posterior probability of colocalization (PPC) was greater than 0.8). An eQTL reflects average gene expression whereas a sQTL represents the joint effect of all splicing phenotypes mapping to a gene. The colocalization analyses were done only in EUR participants owing to GTEx having released summary statistics only for EUR participants.

## RESULTS

### Primary GWAS: chondrocalcinosis

There were two genome-wide significant loci in each of the AFR and EUR cohorts (**Table 1; Supplemental Figure 1**), both on chromosome 6. Locus zoom plots of the loci are presented in **Figure 1**. Strongly supporting that these loci are not false positives, the signals in each ancestral group were at the same genomic location, identified by the closest genes *RNF144B* and *ENPP1*. At *RNF144B* there were different lead SNPs (AFR, rs9396861; EUR, rs1886248), however with very similar effect size (OR_AFR_ = 1.49; OR_EUR_ = 1.44). The locus zoom plots suggested a second genetically-independent locus at RNF144B in EUR (lead SNP rs12524807) which was confirmed by conditioning on rs1886248 (**Figure 1**). The signals at *ENPP1* were different between EUR and AFR (71.5 kb apart), however each signal was entirely within the *ENPP1* gene.

**Figure 1.**
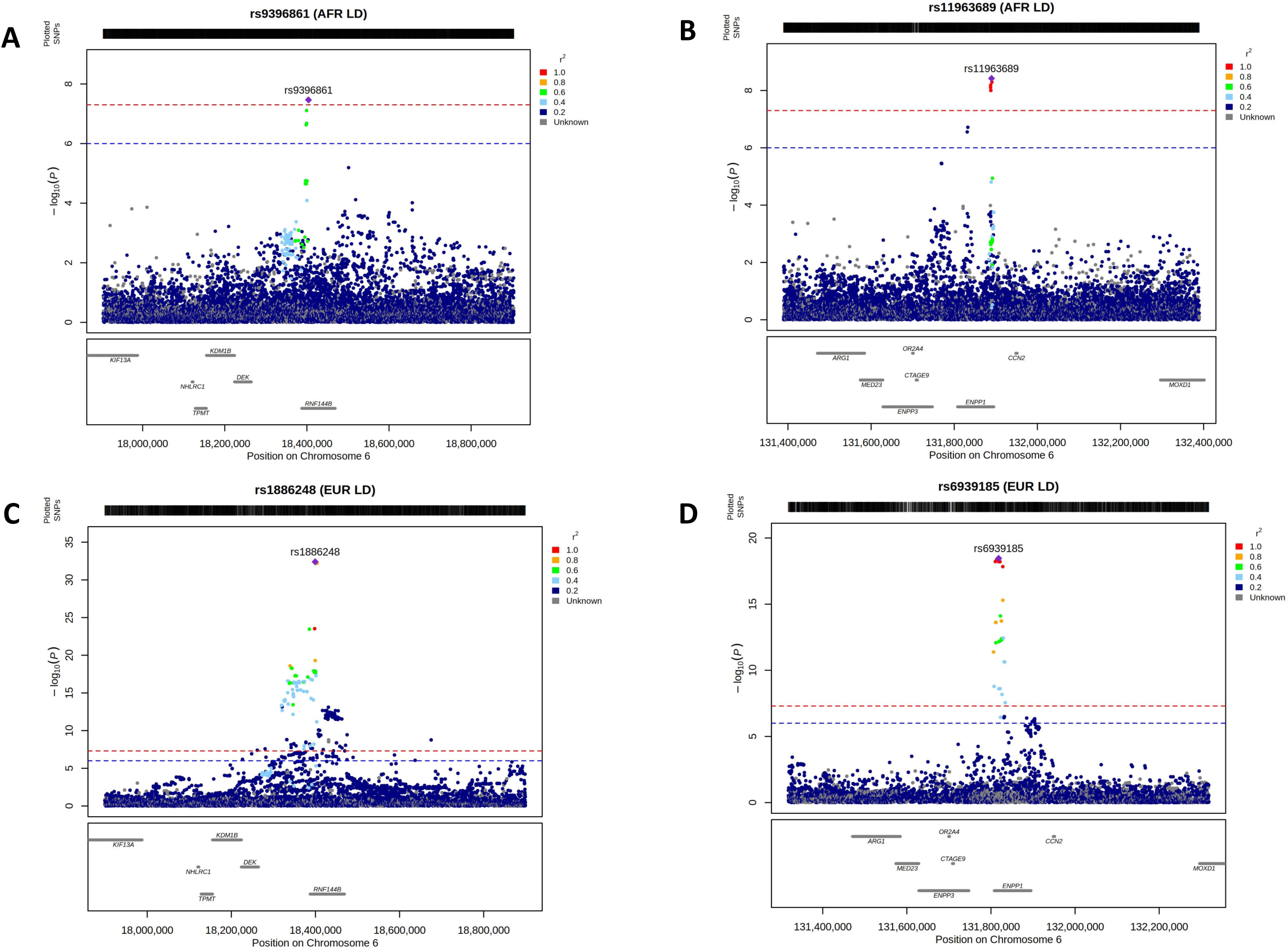
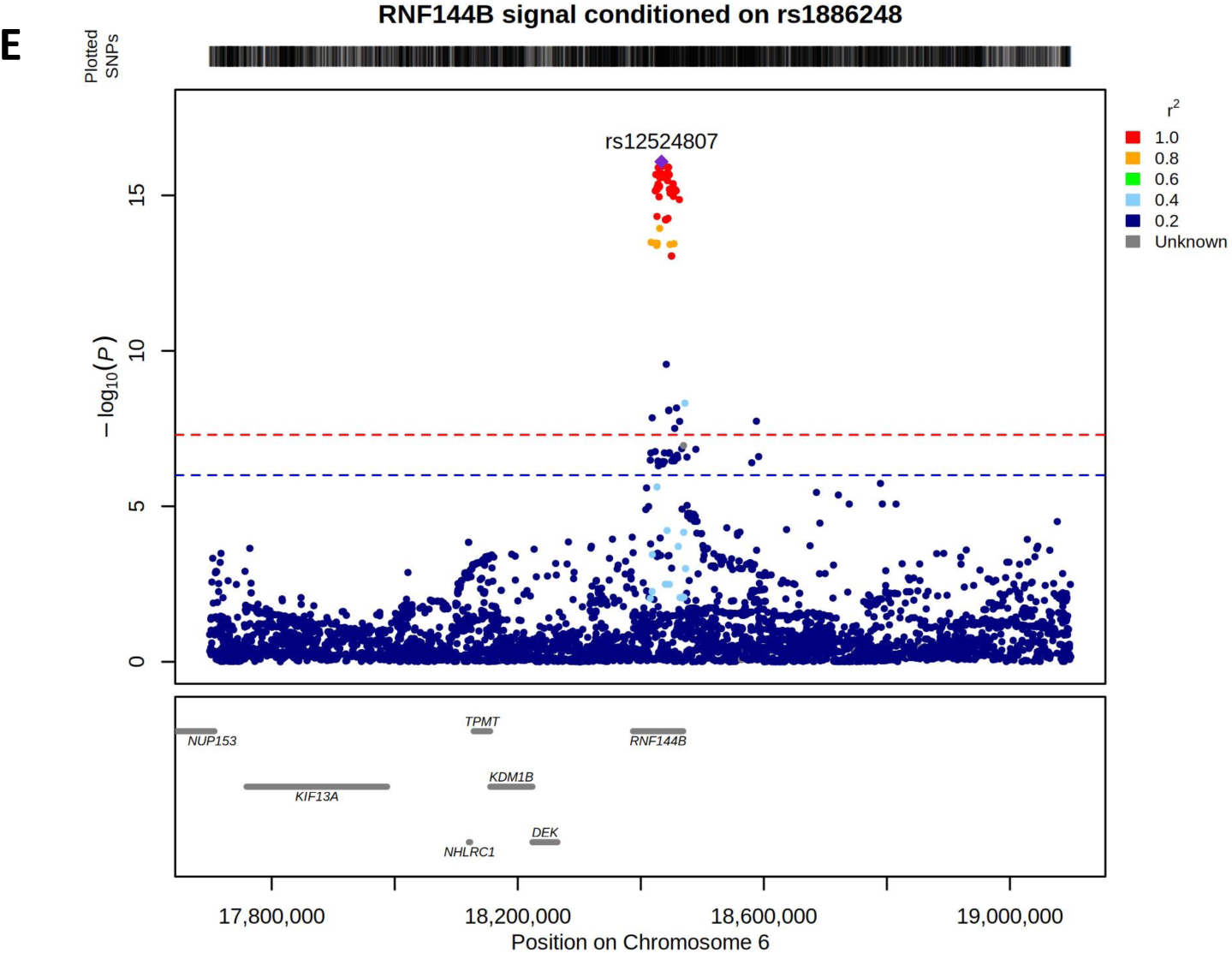
Locus zoom plots of genetic association signals in chondrocalcinosis. A, the *RNF144B* locus in AFR; B, the *ENPP1* locus in AFR; C, the *RNF144B* locus in EUR; D, the *ENPP1* locus in EUR; E, the *RNF144B* locus in EUR after conditioning on rs1886248. Each dot represents a distinct SNP and the extent of linkage disequilibrium with the labelled SNP is indicated by the color key.

The EUR and AFR meta-analysis of chondrocalcinosis did not reveal additional loci that fit our criteria for declaring a disease-associated signal (**Supplementary Table 1**). There were two signals evident in the locus zoom plot of the meta-analyzed *ENPP1* locus that corresponded to each of the EUR and AFR signals (**Figure 2**), albeit with a different lead SNP (rs1409181) at the EUR signal (r^2^ = 0.66 with the EUR rs6939185 lead SNP (**Figure 1**)). At *RNF144B* the lead SNP was rs1886248, the same as in EUR but not as in AFR (**Table 1**, **Figure 2**).

**Figure 2.**
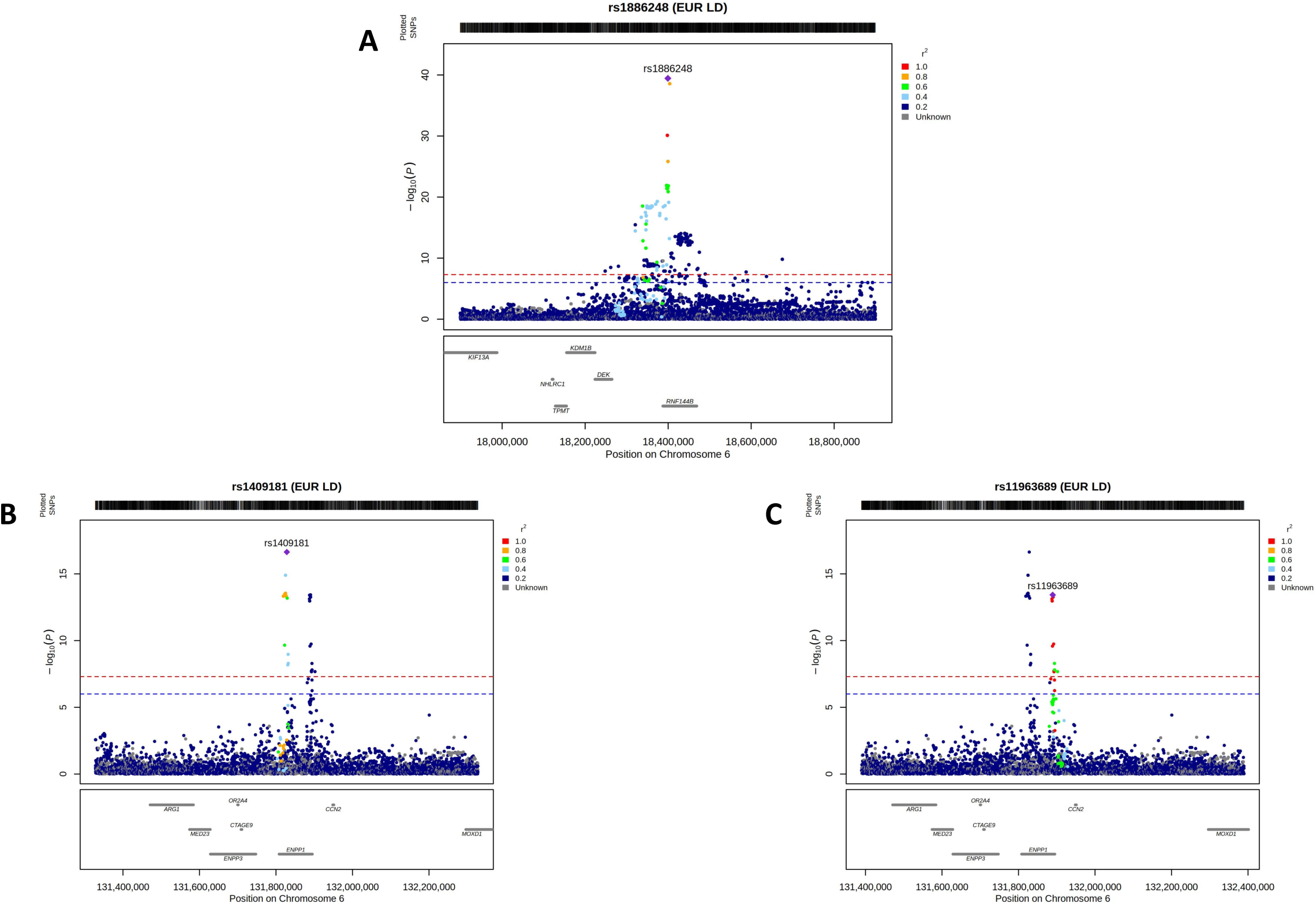
Locus zoom plots of genetic association signals in chondrocalcinosis resulting from meta-analysis of the AFR and EUR GWAS summary statistics. A, the *RNF144B* locus; B, the *ENPP1* locus with the signal attributable to EUR labelled; C, the *ENPP1* locus with the signal attributable to AFR labelled. Each dot represents a distinct SNP and the extent of linkage disequilibrium with the labelled SNP is indicated by the color key.

### Colocalization analysis with gene expression in chondrocalcinosis

Both *RNF144B* and *ENPP1* are widely expressed (**Supplemental Figure 2**). The lead SNPs at the respective loci map within an intron of either *RNF144B* or *ENPP1*, suggesting that their effect on chondrocalcinosis could be mediated through control of gene expression. Therefore, we tested whether the EUR lead variants associate with expression (i.e. are expression quantitative trait loci (eQTL) or alternative splicing quantitative trail loci (sQTL)) in the Genotype and Tissue Expression (GTEx) project^24^. The C (risk) allele of rs1886248 associated with increased expression of *RNF144B* in cultured fibroblasts and skeletal muscle. Furthermore, rs1886248 associated with a *RNF144B* sQTL in 31 tissues with strongest associations in thyroid and whole blood. At the *ENPP1* locus the G (risk) allele of rs6939185 associated with increased expression of *ENPP1* in brain cerebellum, brain cerebellar hemisphere and skeletal muscle. There was no sQTL for rs6939185.

We formally tested whether the chondrocalcinosis genetic association signal genetically colocalized (overlapped) with the genetic signals of association with expression level of *RNF144B* or *ENPP1* and splicing of *RNF144B.* If so, this would suggest two things; one, evidence that the causal gene at the locus was *ENPP1* or *RNF144B* and, two, evidence that control of gene expression and/or alternative splicing mediates the effect of the GWAS signal. The chondrocalcinosis genetic association signal at *ENPP1* genetically colocalized with eQTLs in brain cerebellum and cerebellar hemisphere (posterior probability of colocalization (PPC) = 0.99 and 0.89, respectively) where the G (risk) allele of rs6939185 associates with increased expression of *ENPP1*. However, there was no colocalization with *ENPP1* in skeletal muscle (PPC = 0.00). For *RNF144B* there was no evidence of colocalization of the chondrocalcinosis genetic association signal with an eQTL in cultured fibroblasts or skeletal muscle (PPC = 0.00 and 0.37, respectively). However, in the 31 tissues in which rs1886248 associated with alternative splicing of *RNF144B* there was strong evidence for colocalization of the chondrocalcinosis genetic association signal with genetic control of alternative splicing of *RFN144B* (PPC > 0.91 for all; adipose subcutaneous, adipose visceral omentum, artery aortic, artery coronary, artery tibial, brain cortex, brain putamen basal ganglia, brain anterior cingulate cortex, brain frontal cortex, brain nucleus accumbens basal ganglia, brain hippocampus, brain substantia nigra, brain amygdala, esophagus muscle, esophagus gastroesophageal junction, esophagus mucosa, fibroblasts cultured, heart left ventricle, lung, mammary tissue, muscle skeletal, nerve tibial, ovary, pituitary, prostate, skin sun-exposed, skin not sun-exposed, spleen, uterus, whole blood, thyroid). In summary, this analysis supports both *ENPP1* and *RNF144B* as candidate causal genes for chondrocalcinosis.

We observed in GTEx association of genetic control of gene expression with other genes at *ENPP1*. Specifically, rs6939185 associated with long non-coding RNA *RP1-131F15.2* in thyroid and pituitary, and with *ARG1* in sun-exposed skin. Colocalization was observed with *RP1-131F15.2* in both the thyroid and pituitary (PPC = 0.85 and 0.80, respectively) but not with *ARG1* (PPC = 0.00). The tissue expression pattern of *RP1-131F15.2* largely mirrored that of *ENPP1* (**Supplementary Figure 2**). In summary, this analysis supports *RP1-131F15.2* as a second candidate causal gene for chondrocalcinosis at the *ENPP1* locus.

### Secondary GWAS: crystal arthropathy

The same AFR *ENPP1* chondrocalcinosis lead SNP (rs11963689) was genome-wide significant in AFR crystal arthropathy. However, in EUR there was a different lead SNP (rs766592) at *ENPP1*, which was in strong linkage disequilibrium (r^2^ = 0.97) with the EUR chondrocalcinosis lead SNP (rs6939185) (**Table 1**). At *RNF144B* in EUR the lead SNP was the same as for chondrocalcinosis (rs1886248). The locus zoom plots (not shown) were very similar to those for chondrocalcinosis. The EUR and AFR meta-analysis did not reveal additional loci that fit our criteria for declaring a disease-associated signal.

### Specific investigation of ANKH and chondrocalcinosis

The minor allele (A) of a 5’UTR G>A transition 4 bp upstream of the start codon of the *ANKH* gene (rs78431233) has previously been reported as a risk allele for chondrocalcinosis in two studies among kindreds with early-onset CPPD disease^14,15^. We specifically examined the *ANKH* locus, where there was weak evidence for a signal within the gene in each of EUR (rs2921603, *P* = 3.4 x 10^-4^) and AFR (rs536902008, *P* = 6.0 x 10^-5^) (**Supplemental Figure 3**). However, rs78431233 was only weakly associated with chondrocalcinosis in EUR (*P* = 0.04) although the A allele also conferred risk (OR = 1.14). We also interrogated ClinVar^25^ for clinically relevant genetic variants implicated in chondrocalcinosis (**Supplementary Table 2**). Of the 34 variants included in the AFR-EUR meta-analysis, two variants associated with chondrocalcinosis after correction for multiple testing (*P* < 0.05/34 = 1.5 x 10^-3^; rs17364066 and rs3045). Both SNPs map to the 3’ UTR, and rs3045 is common in AFR and EUR (MAF = 0.11 and 0.09, respectively) whereas rs17364066 is common in EUR (MAF = 0.09) but uncommon in AFR (MAF = 0.004). Both SNPs are in complete linkage disequilibrium in EUR (r^2^ = 1.0).

## DISCUSSION

To our knowledge, this is the first report of a genome-wide association analysis in chondrocalcinosis, which is a radiographic finding typically attributed to CPPD. Illustrating the power of the unbiased approach afforded by GWAS, we identified major signals in separate cohorts of European and African ancestry within the *ENPP1* and *RNF144B* genes. The genetic evidence implicates *ENPP1* as a candidate causal gene mediating the genetic effect; the lead SNPs in each ancestral group mapped within the gene, the chondrocalcinosis risk allele was associated with increased expression of *ENPP1* and the genetic signal of association with chondrocalcinosis colocalized with genetic control of expression. Genetic control of expression of the long non-coding RNA *RP1-131F15.2* also colocalized with the chondrocalcinosis genetic association signal with the risk allele associating with reduced expression of *RP1-131F15.2*. Colocalization of a genetic signal of association with each of a long non-coding RNA and a protein-coding gene is a common phenomenon at GWAS loci (e.g. in gout^26^) and our findings may reflect mediation of regulation of gene expression of *ENPP1* by control of expression of *RP1-131F15.2*, which maps within *ENPP1*. At *RNF144B* our findings suggest that the genetic effect on chondrocalcinosis is mediated by an influence on *RNF144B* transcript isoforms.

*ENPP1* encodes ectonucleotide pyrophosphatase / phosphodiesterase family member 1. It is a transmembrane glycoprotein that hydrolyzes nucleoside triphosphates, primarily adenosine triphosphate (ATP), to produce AMP and PPi. Increased ENPP1 activity, induction of ANK (a regulator of ATP transport) and subsequent production of extracellular inorganic pyrophosphate by chondrocytes could contribute to the formation of calcium pyrophosphate crystals^27-29^. A nucleotide analog (SK4A) was reported as a potent inhibitor of ENPP1 in whole cartilage tissue and could ameliorate ATP-induced calcium pyrophosphate crystal deposition in cultured chondrocytes^30^. Hydrolysis of 2’,3’-cG/AMP by ENPP1 inhibits the innate immune system stimulator of interferon genes (STING) pathway^31^. This has led to the development of selective ENPP1 inhibitors for treatment of infectious disease and cancer^32,33^. In light of the causal link between ENPP1 and chondrocalcinosis, these inhibitors could potentially be repurposed for the treatment, or possibly even prevention, of CPPD in individuals with increased genetic susceptibility. However, this possibility would need careful evaluation as ENPP1 is widely expressed and effective blockage might increase levels of extracellular ATP which can have significant negative effects. Deficiency of ENPP1 is also associated with sometimes severe diseases including congenital ectopic vascular calcification and osteoporosis^34,35^.

RNF144B is a RING domain-containing E3 ubiquitin-protein ligase. It negatively regulates apoptosis^36^, is inducible by lipopolysaccharide in primary human macrophages and promotes IL-1β expression^37^. SNP rs9396861 has previously been associated with finger osteoarthritis by GWAS^38^, with the C-allele increasing risk, consistent with our observation in chondrocalcinosis. How RNF144B could contribute to the pathogenesis of chondrocalcinosis and CPPD disease is unclear.

A limitation of GWAS methodology is that common SNPs do not capture the total heritability of the disease trait of interest. Several factors may contribute to the so-called ‘missing heritability’, including imperfect LD^39^ between SNP marker and causal locus and small marker effect size^40^. These limitations amplify when studying a disease trait with lower heritability. This is the case for chondrocalcinosis (estimated heritability of ∼15% in the AFR cohort used here, and ∼9% in EUR^16^), because the predictive ability of the risk alleles is diminished or sporadic cases of familial disease are governed by rare highly penetrant alleles, which appears to be the case in CPPD disease and related disorders^41^. Nevertheless, first published GWAS of traits typically involve smaller case sample size and can reveal important loci of moderate to large effect with biological significance. For example, the top finding from the early GWAS of serum urate was *SLC2A9^42^* which directly implicated genetic variation in uric acid transporters expressed in the kidney that function to reabsorption and excrete urate. Likewise in the current genetic study of chondrocalcinosis, two moderate effect size loci in *ENPP1* and *RNF144B* have been implicated and suggest pathophysiology of disease. Our study highlights the utility of GWAS in deepening our understanding of genetic mechanisms governing chondrocalcinosis and CPPD disease.

There are several limitations to this study. First, crystal arthropathy may not necessarily be due to CPPD disease, and could potentially include misclassified gout. Whilst we are unable to determine the extent to which this might have occurred, as these were publicly available GWAS summary statistics, we are confident that the crystal arthropathy Phecode accurately captures CPPD disease, given that we detected no signal at the strong effect gout loci *SLC2A9* and *ABCG2^26^* (**Supplemental Figure 4**). Furthermore, the Phecode for calcium pyrophosphate deposition (chondrocalcinosis) used by the MVP^16^ has been validated in the Veterans Affairs system^43^. Second, chondrocalcinosis may be under-ascertained in the controls if they did not have a radiograph performed and/or coded, however this would bias the results towards the null. Thirdly, an automated approach had been used for phenotyping^16^, which is efficient but has not been validated using medical record review for chondrocalcinosis or crystal arthropathy. Mitigating the possibility of misclassification of other conditions as chondrocalcinosis or crystal arthropathy, Phecodes are derived using ICD codes and a prior study using VA data reported that CPPD was accurately identified using a single ICD-9 code^43^. Finally, investigation using GTEx of association of the chondrocalcinosis genetic association signal with genetic control of gene expression and alternative splicing is limited by the tissues with gene expression data available. For example there are no data for cartilage, and we were unable to analyze genetic control of gene expression in AFR in GTEx using the publicly available GTEx data.

## Data Availability

All data produced in the present work are contained in the manuscript

http://gtexportal.org

## Acknowledgements

We thank MVP participants for their service and their continued contributions to the United States through participation in this study. We thank the Million Veteran Program, Office of Research and Development, and Veterans Health Administration for supporting the work in ref^16^ which generated the summary statistics on which our work was based. This work was supported in part by Merit Review I01 BX004454 from the United States Department of Veteran’s Affairs (to AR) and in part by Rheumatology Research Foundation R Bridge (to SKT).

## Figure legends

**Supplementary Figure 1.** Manhattan plots. A, chondrocalcinosis in AFR; B, crystal arthropathy in AFR; C, chondrocalcinosis in EUR; D, crystal arthropathy in EUR; E, chondrocalcinosis in AFR+EUR; F, crystal arthropathy in AFR+EUR. The red line is the threshold for genome-wide significance (*P* = 5×10^-8^) and the blue line is the threshold for suggestive significance (*P* = 1×10^-6^).

**Supplementary Figure 2.** Bulk tissue gene expression of *RNF144B* (A), *ENPP1* (B) and *RP1-131F15.2* (C) in GTEx data.

**Supplementary Figure 3.** Locus zoom plots of the *ANKH* locus in AFR (left) and EUR (right). Each dot represents a distinct SNP and the extent of linkage disequilibrium with the labelled SNP is indicated by the color key.

**Supplementary Figure 4.** Locus zoom plots of the strongly gout-associated loci *ABCG2* (left) and *SLC2A9* (right) in EUR crystal arthropathy. Each dot represents a distinct SNP and the extent of linkage disequilibrium with the labelled SNP is indicated by the color key.

## Footnotes

^ The contents of this manuscript do not represent the views of the US Department of Veteran’s Affairs or the United States Government.

