## Supplementary figures and images for "Genome-wide association study in chondrocalcinosis reveals ENPP1 as a candidate therapeutic target in calcium pyrophosphate deposition disease"

### Supplementary Figures

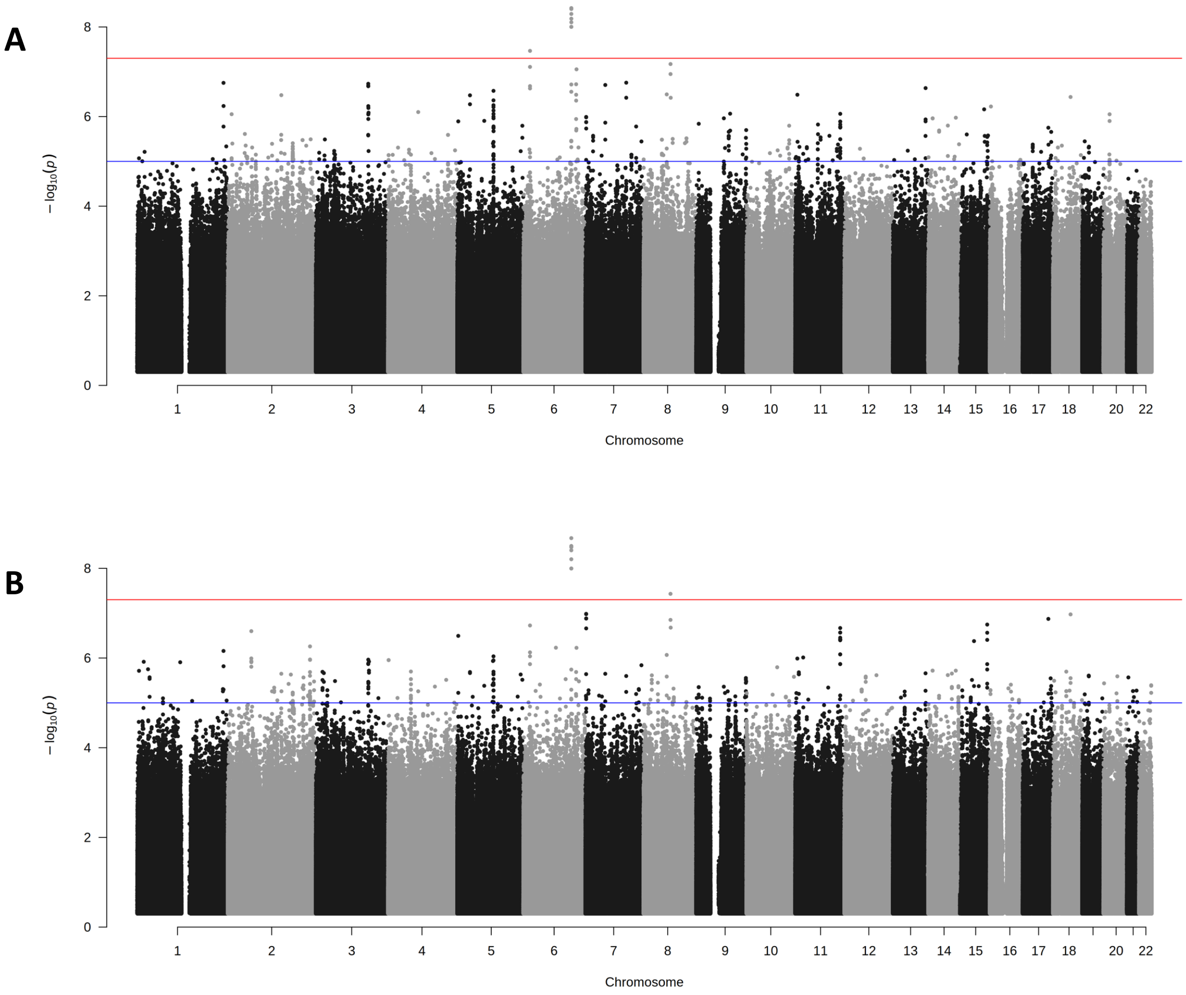

**Figure S1 cont.**

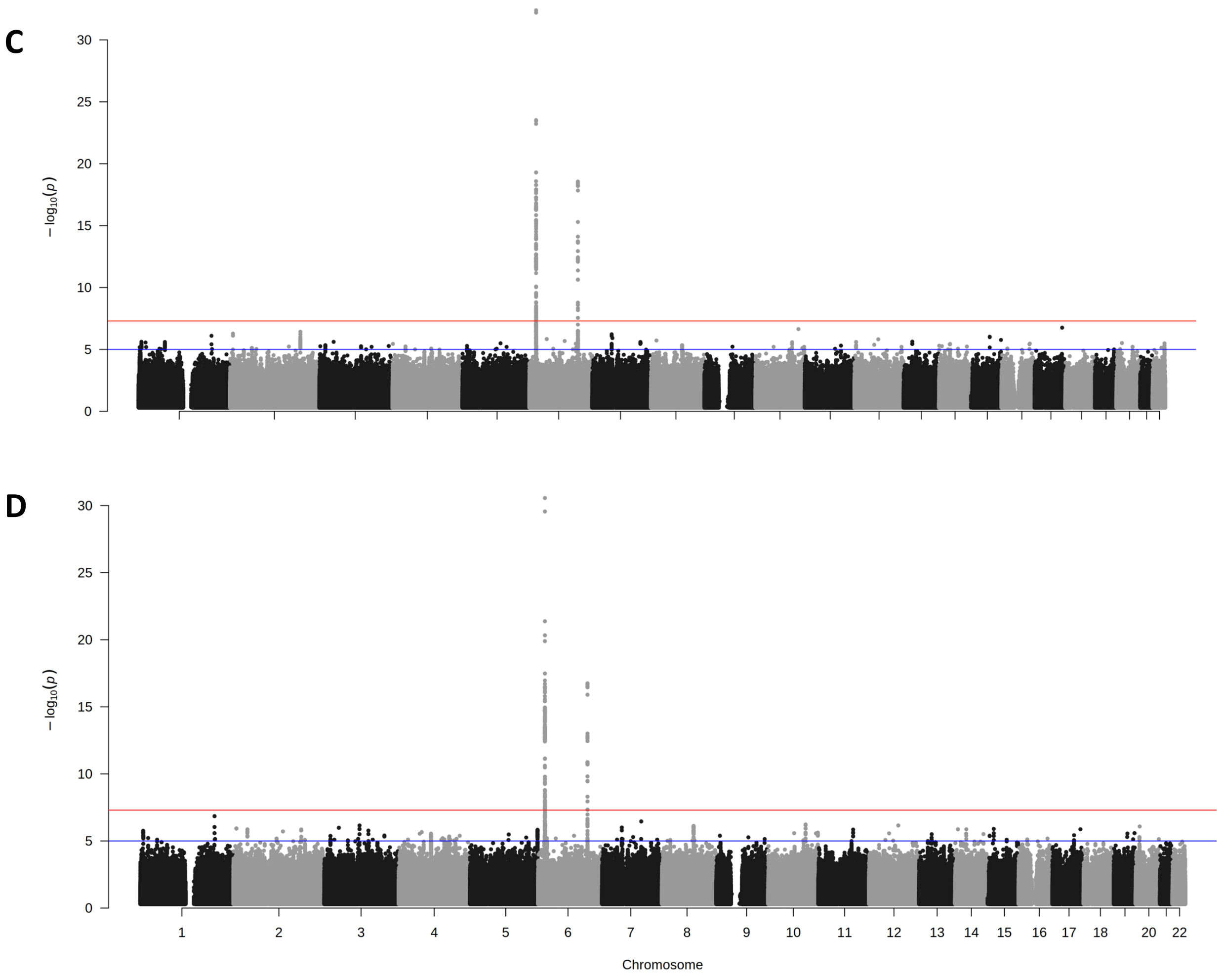

**Figure S1 cont.**

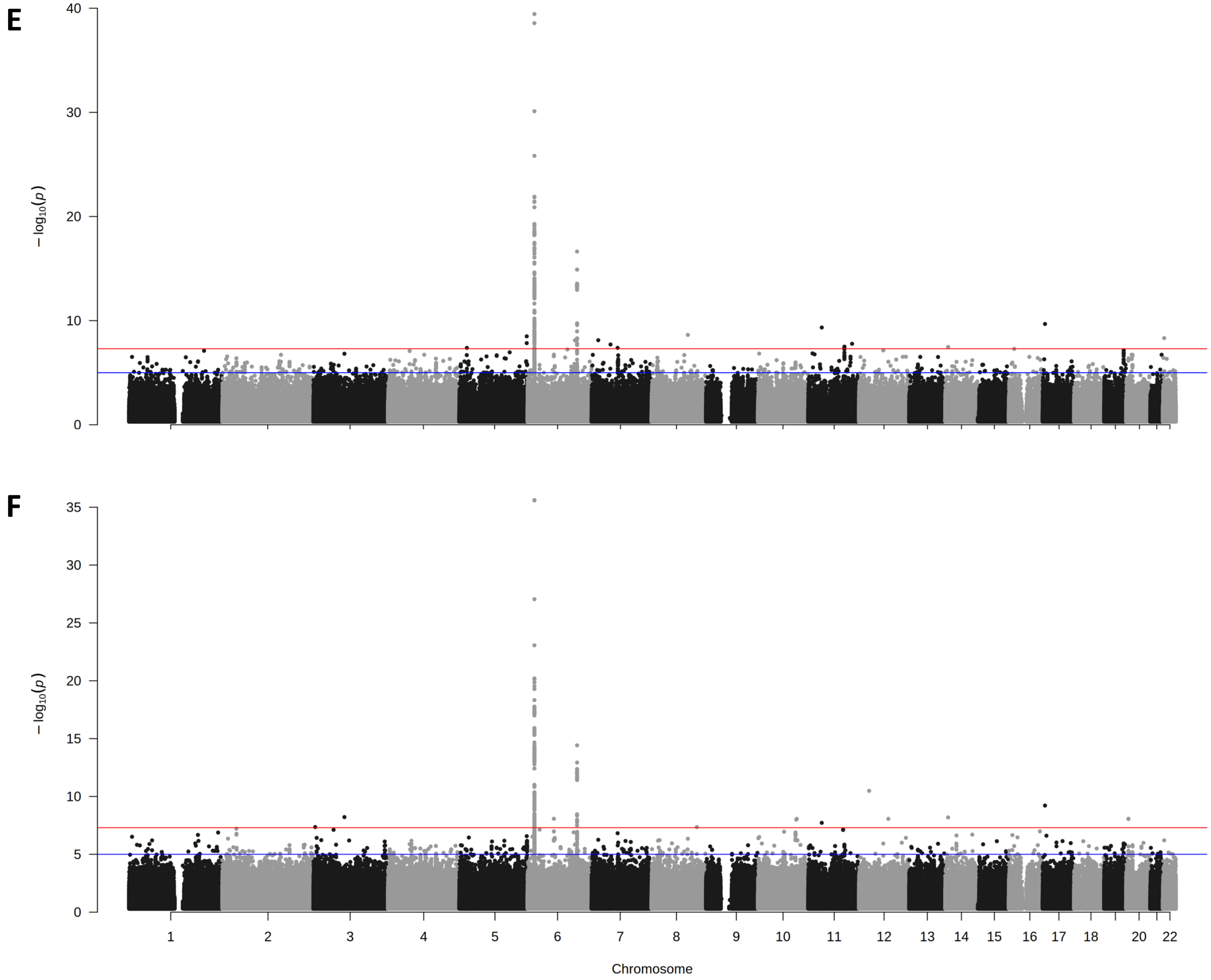

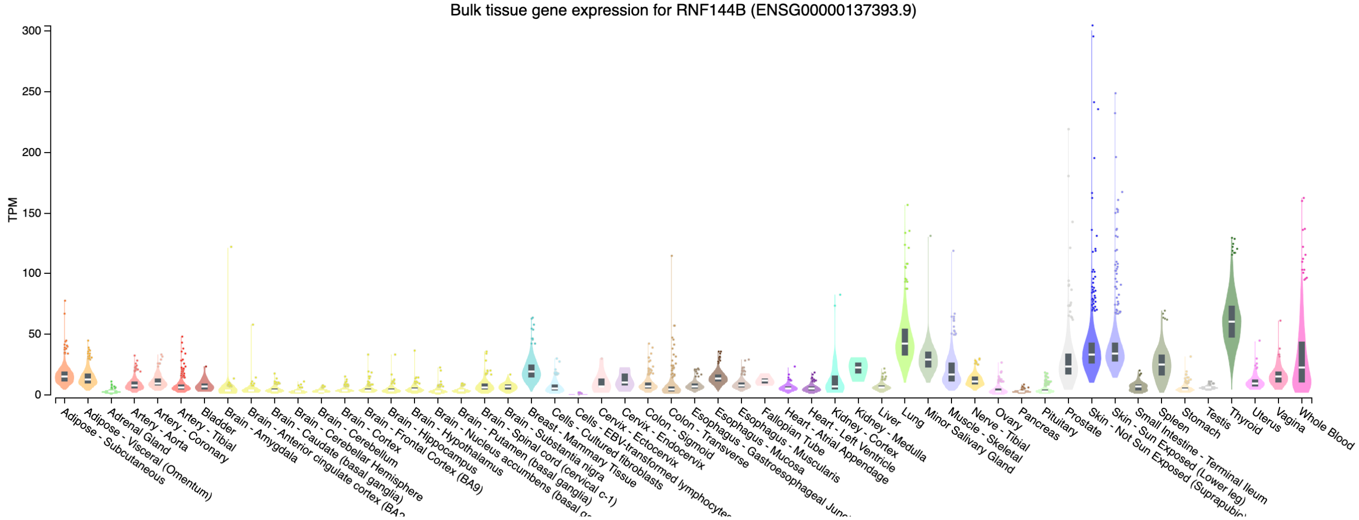

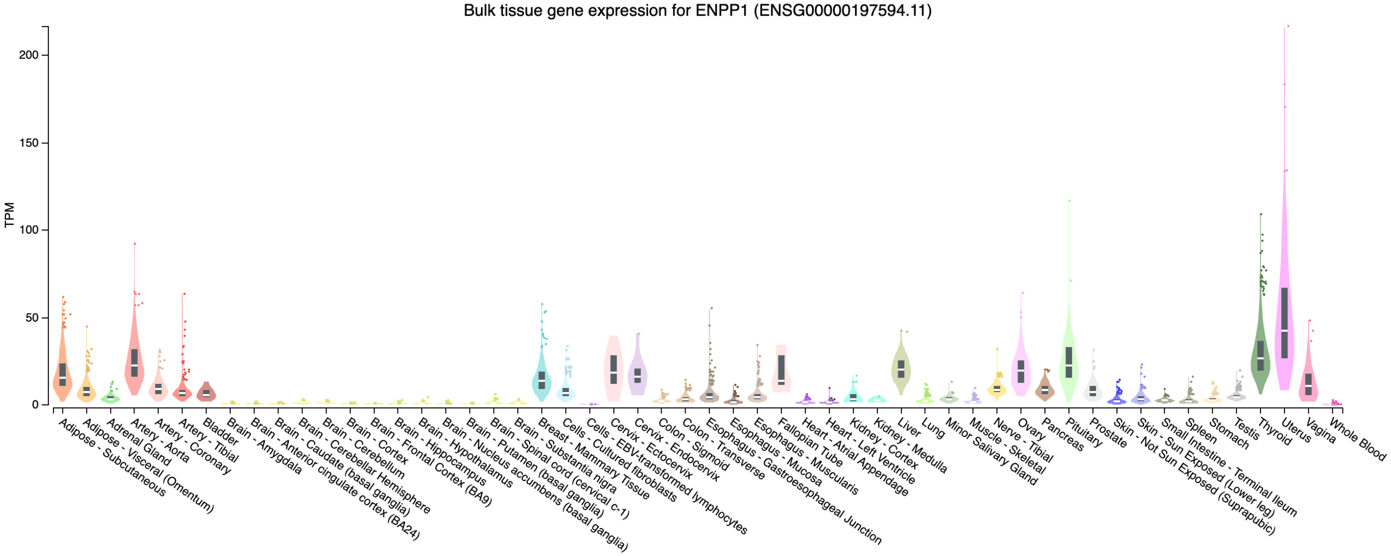

#### Figure S2 cont.

C

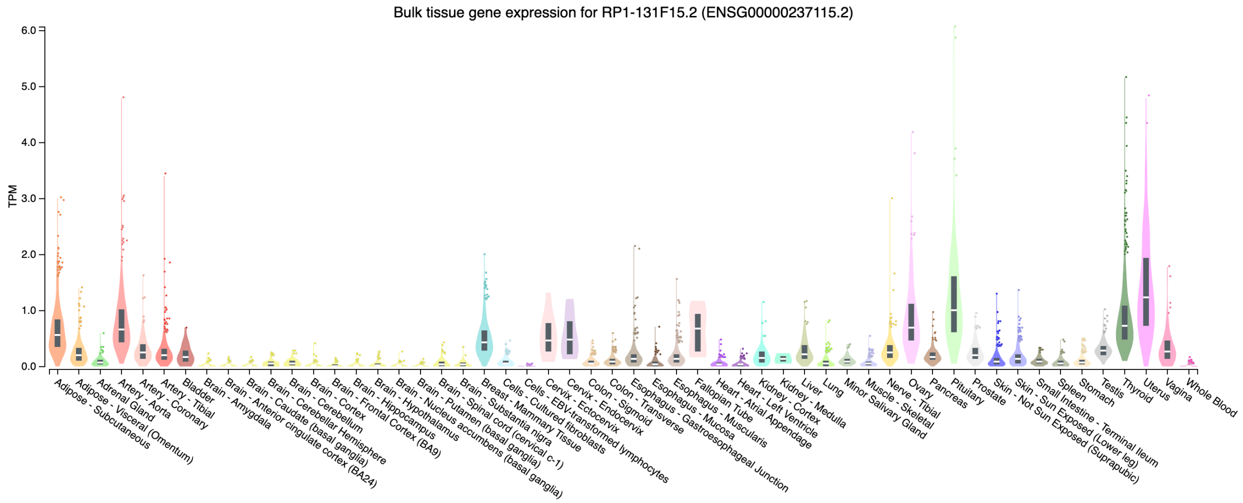

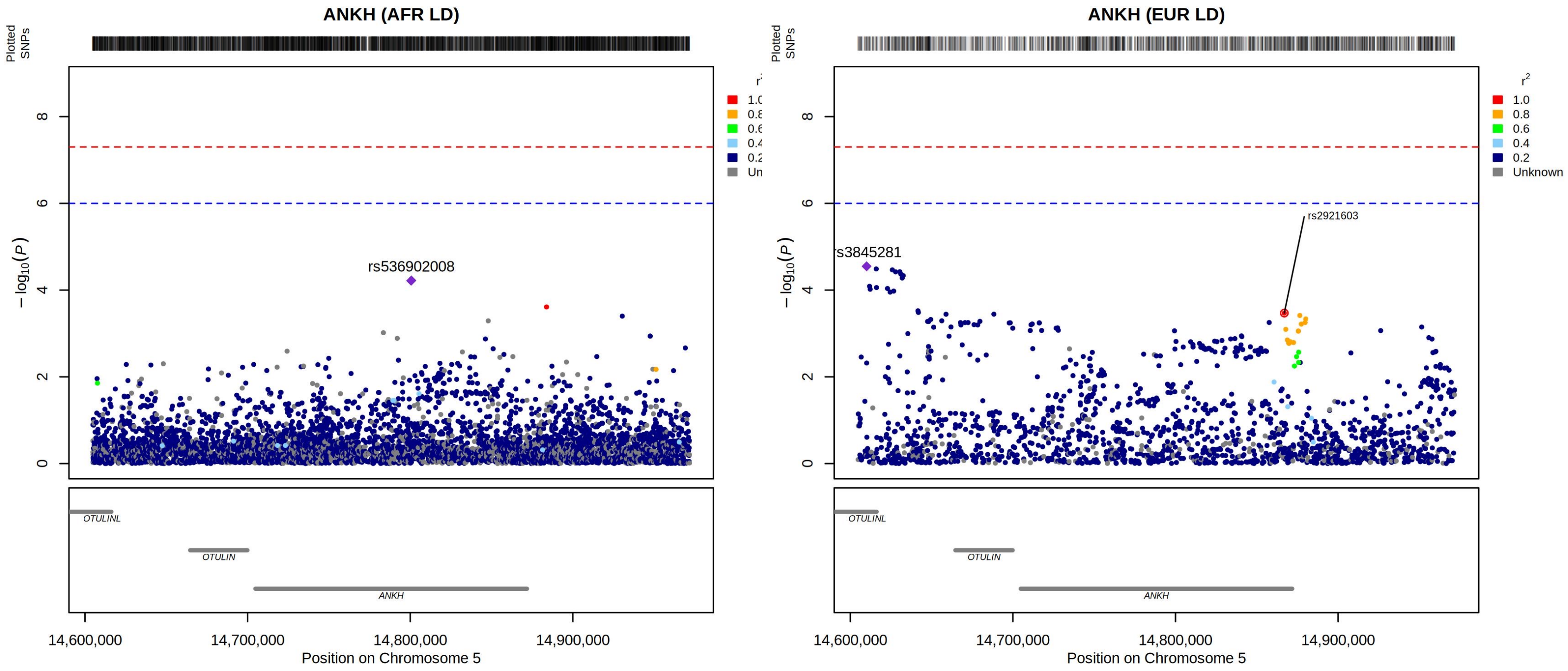

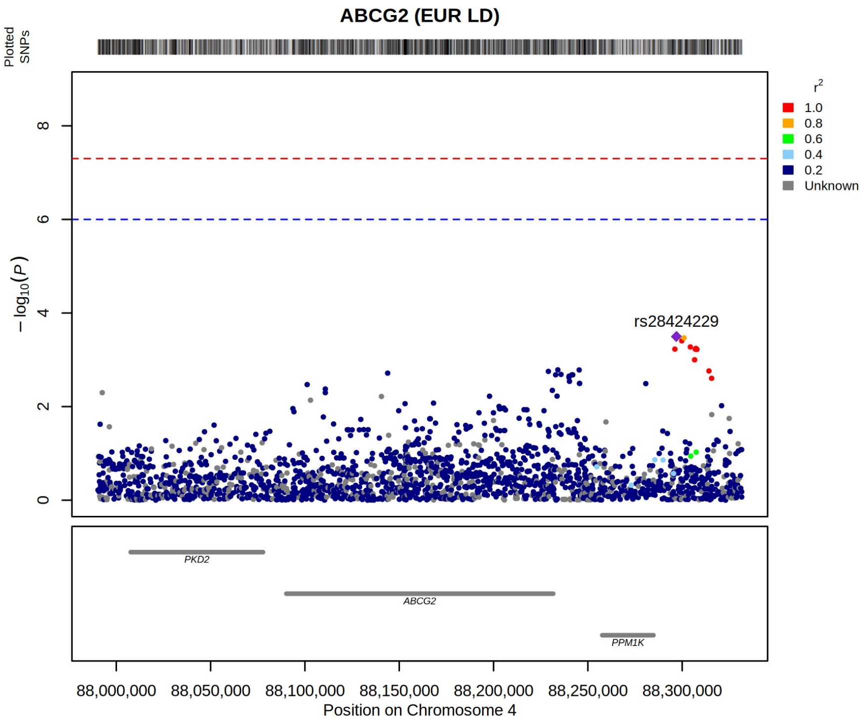

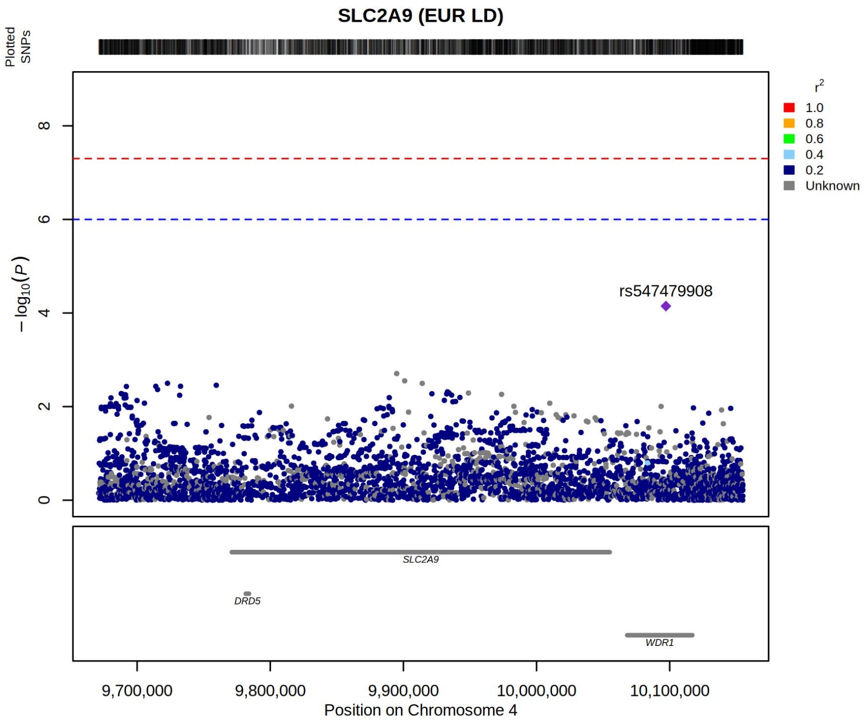
